# Experience in the implementation of the early warning sign system in obstetric patients (Maternal Early Warning Criteria, MEWC) during early postpartum. A prospective observational study

**DOI:** 10.1101/2020.09.15.20194910

**Authors:** Cristina Ibáñez-Lorente, Rubén Casans-Francés, Soledad Bellas-Cotán, Luis E. Muñoz-Alameda

## Abstract

**Objective:** To evaluate the implementation of an early warning system in obstetric patients (MEWC) during the first two hours after delivery in a single tertiary-care hospital.

**Methods:** The MEWC system implementation was carried out from 15th March to 15th September 2018, over 1166 patients. The parameters collected were systolic and diastolic blood pressure, heart rate, oxygen saturation, diuresis, uterine involution, and bleeding. If a parameter was not within defined limits, an obstetrician first examined the patient, determining the need to call the anesthesiologist. We carried out a sensitivity-specificity study of the trigger and multivariate analysis of the factors involved in developing potentially fatal disorders (PFD), reintervention, critical care admission, and stay.

**Results:** The protocol was triggered in 75 patients (6.43%). The leading cause of alarm activation was the altered systolic blood pressure (32 [42.7%] patients), and eleven developed PFD. Twenty-eight patients were false-negatives. Sensitivity and specificity of MEWC protocol were 0.28 (0.15, 0.45) and 0.94 (0.93, 0.96), respectively. Multivariate analysis showed a relationship between alarm activation and PFD.

**Conclusion:** Our MEWC protocol presented low sensitivity and high specificity, having a significant number of false-negative patients.

## Introduction

Lowering morbidity and mortality in the obstetric patient continues to be a quality-of-care criterion for healthcare centers. The development over the few decades shows a remarkable improvement in healthcare indicators in this population group [1]. Although mortality rates are very low for developed countries, the impact is high, both for the social repercussion that it involves and the years of life lost.

One of the approaches to reduce maternal morbidity and mortality has been using tools that would enable the rapid identification of patients who would benefit most from an aggressive intervention or a higher level of care. In December 2007, a review on maternal mortality concluded that 40-50% of the UK’s mortality rates are preventable. However, the early warning signs were often not recognized [2]. Since then, most UK hospitals have adopted Maternity Early Obstetric Warning Criteria (MEWS) protocols, with these being an auditable quality criterion for the centres [3, 4].

Unfortunately, the implementation of these protocols is not universal, and its use is not common practice outside English-speaking countries. Furthermore, although the adoption of these protocols has shown an improvement in the quality of assistance to pregnant women and the detection of adverse effects, it has not shown any impact on reducing maternal mortality [5].

This study evaluates the usefulness and feasibility of implementing a modified obstetric early warning system in our center and studying its possible relationship with the detection and identification of adverse effects.

## Methods

### Study design and setting

This study analyses the implementation of a MEWC protocol in a tertiary-care hospital (Hospital Fundación Jiménez Díaz, Madrid, Spain). The hospital has all the medical and surgical specialties but does not have a specific building for gynecology and obstetrics. We designed a single-arm prospective cohort study developed during the first six months after implementing the protocol (first day of implementation: 15th March 2018). Our center’s ethics committee approved our research before the start of patient recruitment (Date of Approval: 12th March 2018, Code: FJD-MEOWS-17-01. S1 File, original in Spanish). This study followed the Strengthening the Reporting of Observational Studies in Epidemiology (STROBE) reporting guideline for cohort studies [6].

### Sample size calculation

For calculating the sample size [7], we considered that our model should have a minimum sensitivity and specificity of 90%. We knew that the incidence of maternal morbidity in our environment was 5% [8]. Thus, the required sample size with a precision of 0.1, a type I error of 0.05, and a dropout rate of 10% were 692 patients. Since the number of births in the hospital was around one thousand between 1st September 2017 and 28th February 2018, we decided to include all patients who gave birth between 15th March and 15th September 2018, except those who refused to provide their data to the study after informed consent.

### Procedures

We designed our center’s MEWC protocol based on the one described by Mhyre et al. [9]. We monitored systolic and diastolic blood pressure, heart rate, oxygen saturation, and diuresis at 10, 30, 60, 90, and 120 minutes. By consensus between anesthesia and obstetrics departments, we included uterine involution measurement (by manual exploration), diuresis, and any bleeding greater than 500 ml in the protocol. We defined bleeding or lack of uterine involution as obstetric causes. S2 File shows the data collection form.

Two months before the start of implementation, we provided training sessions on the usefulness of the MEWC protocols and the process of implementing them in our hospital to professionals from both services, including midwives. After implementing the protocol, nurses and midwives monitored patients for 2 hours after giving birth or undergoing a cesarean delivery by filling in the data form, either in the delivery room or in the PACU. They also collected data on previous parity, preeclampsia, and multiple births as possible risk factors. For each variable in the protocol, specific values obliged the midwives to call the obstetricians, who had to assess the patient within 15 minutes. If the obstetricians did not resolve the alarm, they would make a second call to the anesthesiologists. Independent of the form’s data collection, the anesthesiologists followed all patients who had given birth until discharge.

### Outcomes

Following the recommendations made by the WHO [10], the outcome of our study was the rate of potential fatal disorder (PFD) during the stay, defined as one of the following criteria: access to a critical care unit (CCU), surgery within two hours of delivery, or length of stay of more than seven days depending on the activation of the alert [11]. As secondary outcomes, we measured the alert activation relation on each of the PFD criteria separately.

### Statistical analysis

We used R v4.0.2 (The R Foundation for Statistical Computing, Vienna, Austria) and RStudio 1.2.5033 (RStudio PBC, Boston, USA) to perform statistical analysis. We analyzed outcomes depending on the alarm activation. We described discrete and continuous variables as number and percentage and median (interquartile range [IQR]), and their differences analyzed using the Pearson test or the Wilcoxon rank-sum tests. We calculated the sensitivity, specificity, positive and negative predictive value (PPV and NPV) of alert activation for PFD and each criterion separately, with a 95% confidence interval. We performed a multivariate logistic analysis to study the association of the outcomes with alarm activation, preeclampsia, parity, type of delivery, and multiple births. We presented the results in forest plots as an odds ratio with a 95% confidence interval. We used Cox regression for multivariate analysis of length of stay, showing the results in forest plot as a hazard ratio with a 95% confidence interval. To avoid errors by multiple comparisons, we calculated the respective q-value for each p-value to maintain a false discovery rate below 5% [12]. We considered comparisons in which the p-value and q-value were below .05 as being statistically significant. We provide the original study databases, the step-by-step statistical analysis, and the document in R-Markdown format in S3 File, S4 File, and S5 File.

## Results

During the study period, there were 1169 deliveries at our hospital. Only three patients declined to participate in the study. Since there were no losses to follow-up, we included 1166 patients. Fig 1 shows the STROBE flow chart.

**Fig 1.**
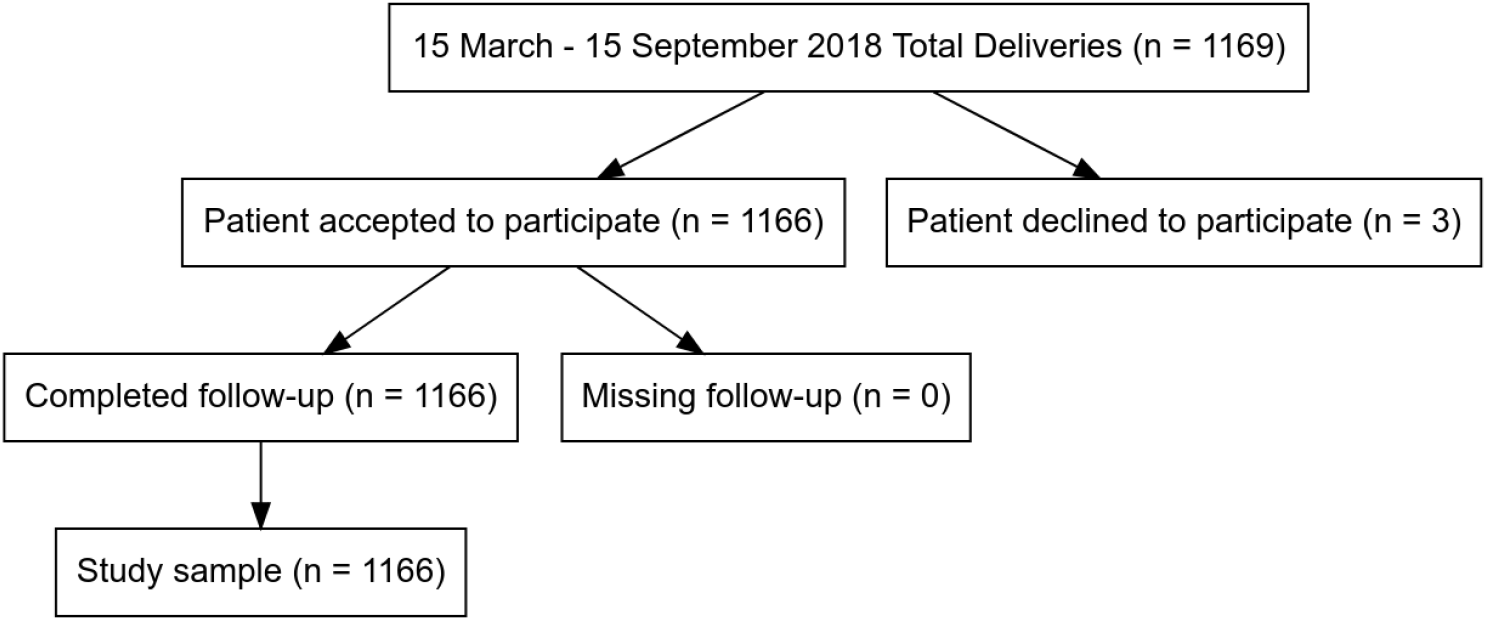
Patients Flowchart according to the STROBE statement.

The patients had a median age of 34 (31-37) years, without previous deliveries, and had a rate of cesarean delivery and instrumental delivery of 23.2% and 15.1%, respectively. The protocol alarm was activated in 75 patients (6.43%). The cohort of subjects with an activated alarm had a higher rate of cesarean delivery (37.3% vs 22.3%, p = 0.005), preeclampsia (17.3% vs 4.8%, p ¡ 0.001), and multiple birth (6.7% vs 1.5%). Also, the PFD rate was higher (14.7% vs 2.6%, p ¡ 0.001), as was the CCU admission rate (12% vs 0.8%, p ¡ 0.001) and length of stay (median: 2.9 [2.3-3.6 vs 2.5 [2.1-3.1] days, p = 0.005). Table 1 shows the demographic data and the outcome results according to MEWC protocol activation.

**Table 1.**
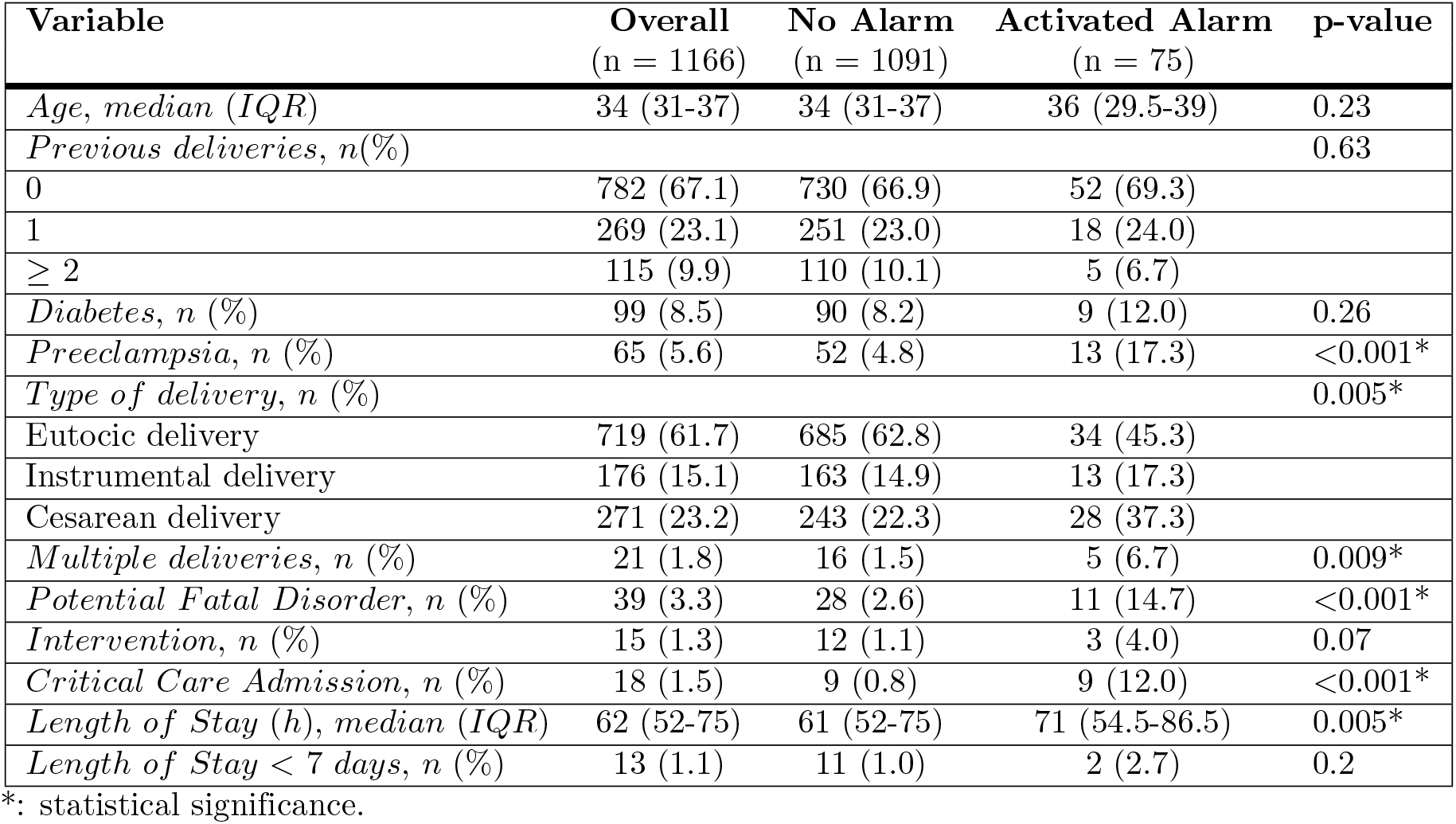
Demographic data and outcome results according to MEWC protocol alarm activation.

| Variable | Overall<br>(n = 1166) | No Alarm<br>(n = 1091) | Activated Alarm<br>(n = 75) | p-value |
| --- | --- | --- | --- | --- |
| Age, median (IQR) | 34 (31-37) | 34 (31-37) | 36 (29.5-39) | 0.23 |
| Previous deliveries, n(%) |  |  |  | 0.63 |
| 0 | 782 (67.1) | 730 (66.9) | 52 (69.3) |  |
| 1 | 269 (23.1) | 251 (23.0) | 18 (24.0) |  |
| $\geq 2$ | 115 (9.9) | 110 (10.1) | 5 (6.7) | |
| Diabetes, n (%) | 99 (8.5) | 90 (8.2) | 9 (12.0) | 0.26 |
| Preeclampsia, n (%) | 65 (5.6) | 52 (4.8) | 13 (17.3) | $<0.001^*$ |
| Type of delivery, n (%) |  |  |  | 0.005* |
| Eutocic delivery | 719 (61.7) | 685 (62.8) | 34 (45.3) |  |
| Instrumental delivery | 176 (15.1) | 163 (14.9) | 13 (17.3) |  |
| Cesarean delivery | 271 (23.2) | 243 (22.3) | 28 (37.3) |  |
| Multiple deliveries, n (%) | 21 (1.8) | 16 (1.5) | 5 (6.7) | 0.009* |
| Potential Fatal Disorder, n (%) | 39 (3.3) | 28 (2.6) | 11 (14.7) | $<0.001^*$ |
| Intervention, n (%) | 15 (1.3) | 12 (1.1) | 3 (4.0) | 0.07 |
| Critical Care Admission, n (%) | 18 (1.5) | 9 (0.8) | 9 (12.0) | $<0.001^*$ |
| Length of Stay (h), median (IQR) | 62 (52-75) | 61 (52-75) | 71 (54.5-86.5) | 0.005* |
| Length of Stay < 7 days, n (%) | 13 (1.1) | 11 (1.0) | 2 (2.7) | 0.2 |

\*: statistical significance.

The leading cause of alarm activation was altered systolic blood pressure (32 [42.7%] patients), followed by obstetric causes (24 [32%] patients). The median time of alarm activation, obstetric assessment, call to the anesthesiologist, and anesthetic assessment was 10, 11, 15, and 20 minutes from the start of postpartum monitoring (Table 2).

**Table 2.** Causes for activation and limits of the MEWC protocol’s trigger thresholds and assessment time from patient monitoring.

| Variable | Activated Alarm (n = 75) | Lower boundary | Upper boundary |
| --- | --- | --- | --- |
| Alarm trigger, n (%) |  |  |  |
| Oxygen saturation | 2 (2.7) | 95% |  |
| Systolic arterial pressure | 32 (42.7) | 90 mmHg | 160 mmHg |
| Diastolic arterial pressure | 3 (4.0) | 40 mmHg | 100 mmHg |
| Heart rate | 14 (18.7) | 50 bpm | 120 bpm |
| Obstetrics (uterine involution and bleeding) | 24 (32.0) | 500 ml |  |
| Urine output | 0 (0) | 17.5 ml/h |  |
| Time to activate the alarm (min), median (IQR) | 10 (0-60) |  |  |
| Time to obstetrics assessment (min), median (IQR) | 11 (2.5-60) |  |  |
| Time to call the anesthesiologist (min) | 15 (4.5, 61.5) |  |  |
| Time to anesthetic assessment (min) | 20 (7-65.5) |  |  |

Our MEWC had a sensitivity of 0.28 (0.15, 0.45), a specificity of 0.94 (0.93, 0.96), a positive predictive value of 0.15 (0.08-0.25) and a negative predictive value of 0.97 (0.96, 0.98) for PFD detection (Table 3). Furthermore, the protocol showed a specificity of 0.94 and a negative predictive value of 0.99 for each of the PFD criteria separately (intervention, CCU admission, and length of stay longer than seven days).

**Table 3.**
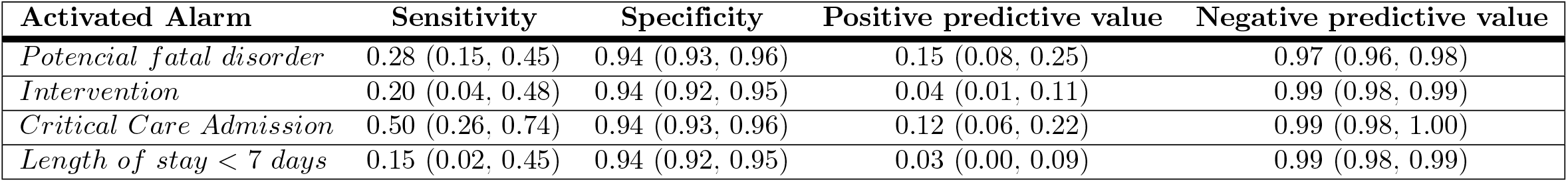
Sensitivity, specificity, positive and negative predictive values (with 95% confidence interval) presented by the activation of our MEWC protocol alarm for the detection of PFD and its respective criteria.

| Activated Alarm | Sensitivity | Specificity | Positive predictive value | Negative predictive value |
| --- | --- | --- | --- | --- |
| Potencial fatal disorder | 0.28 (0.15, 0.45) | 0.94 (0.93, 0.96) | 0.15 (0.08, 0.25) | 0.97 (0.96, 0.98) |
| Intervention | 0.20 (0.04, 0.48) | 0.94 (0.92, 0.95) | 0.04 (0.01, 0.11) | 0.99 (0.98, 0.99) |
| Critical Care Admission | 0.50 (0.26, 0.74) | 0.94 (0.93, 0.96) | 0.12 (0.06, 0.22) | 0.99 (0.98, 1.00) |
| Length of stay < 7 days | 0.15 (0.02, 0.45) | 0.94 (0.92, 0.95) | 0.03 (0.00, 0.09) | 0.99 (0.98, 0.99) |

With regard to multivariate regression analysis, we only found a relationship with PFD with 2 factors, preeclampsia [Odds Ratio = 7.81 (3.50, 16.7), p < 0.001] and MEWC alarm activation [Odds Ratio = 3.97 (1.63, 8.95), p = 0.001] (Fig 2). The only factor related to the intervention was the MEWC alarm activation (Odds Ratio = 4.73 [1.04, 15.7], p = 0.02, Fig 3). The main factor related to critical care admission was preeclampsia (Odds Ratio = 23.2 [7.75-75.3], p < 0.001), followed by MEWC alarm activation (Odds Ratio = 9.73 [2.98-31.5], p < 0.001) and cesarean delivery (Odds Ratio = 4.40 [1.22-18], p = 0.03) (Fig 4). We found no relationship between length of stay and MEWC alarm activation in the Cox regression (Fig 5).

**Fig 2.**
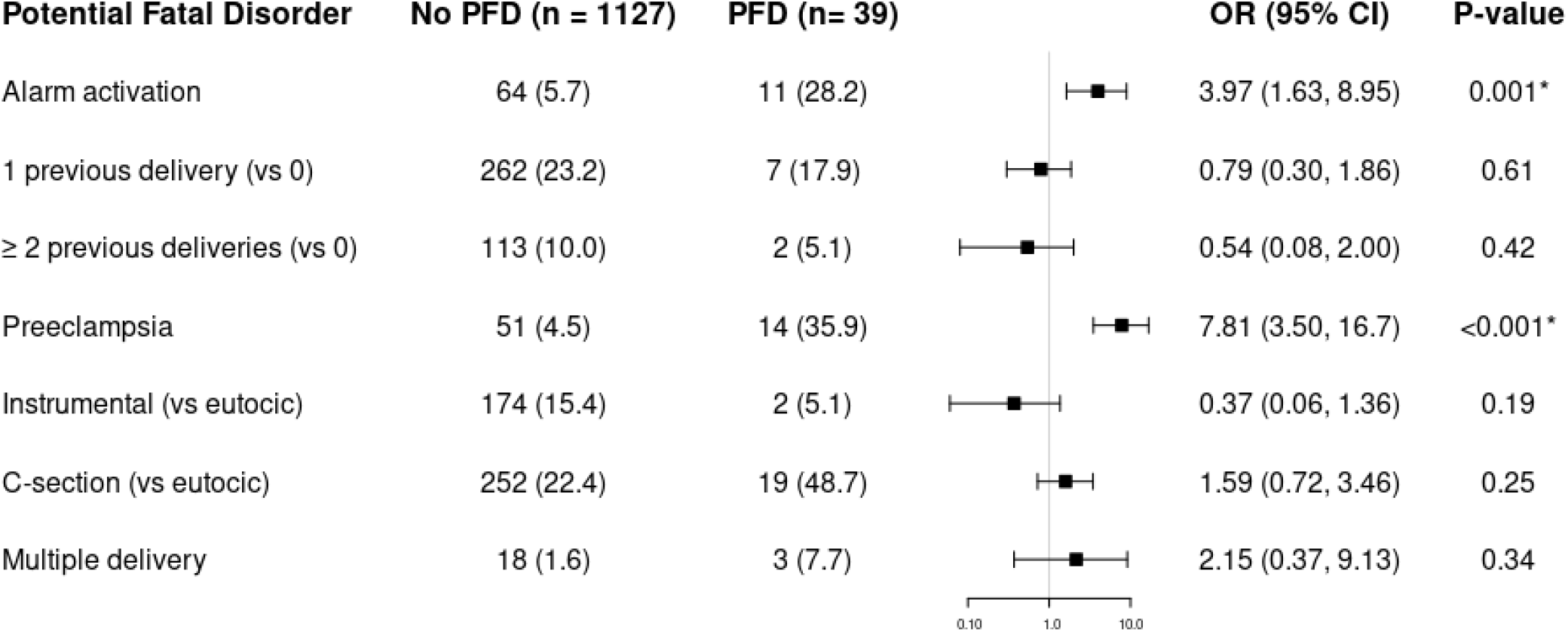
Forest plot of multivariate logistic analysis of the influence of MEWC activation, patient comorbidity and delivery on PFD. We present the results as an odds ratio with a 95% confidence interval. Results less than 1, left of the y-axis, imply risk reduction. We accepted p < 0.05 as significant.

**Fig 3.**
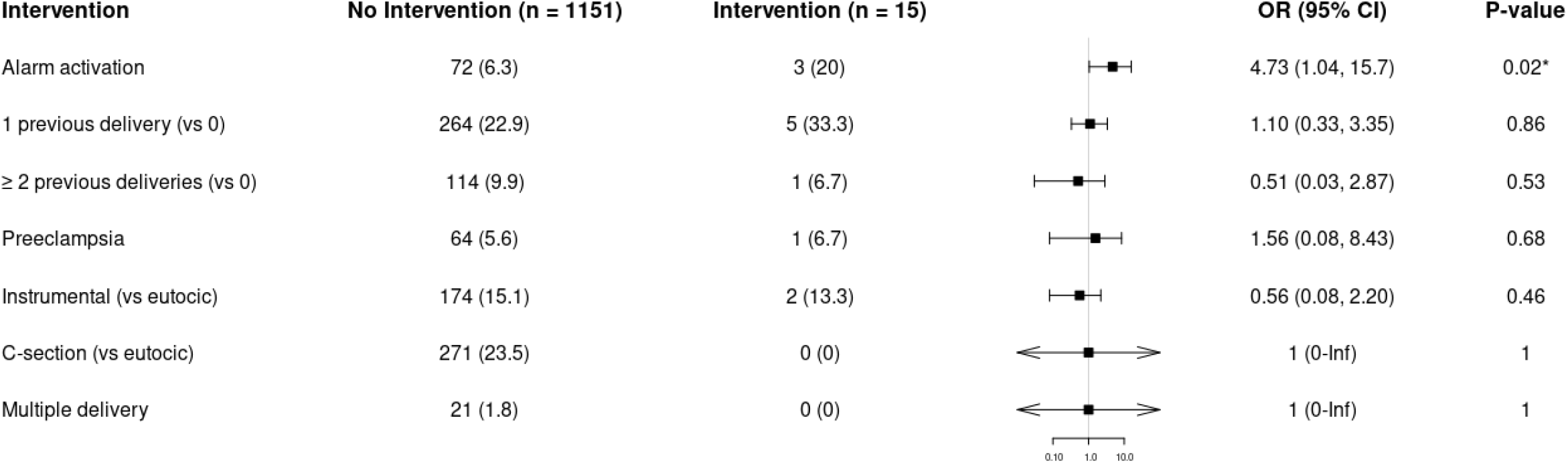
Forest plot of multivariate logistic analysis of the influence of MEWC activation, patient comorbidity and delivery on intervention. We present the results as an odds ratio with a 95% confidence interval. Results less than 1, left of the y-axis, imply risk reduction. We accepted p < 0.05 as significant.

**Fig 4.**
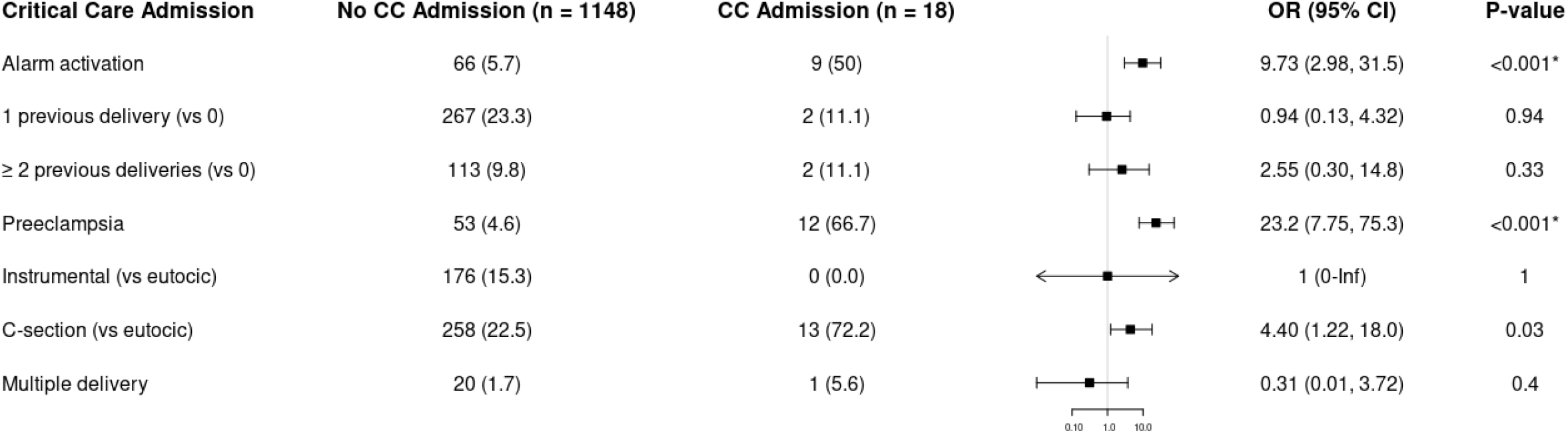
Forest plot of multivariate logistic analysis of the influence of MEWC activation, patient comorbidity and delivery on critical care unit admission. We present the results as an odds ratio with a 95% confidence interval. Results less than 1, left of the y-axis, imply risk reduction. We accepted p < 0.05 as significant.

**Fig 5.**
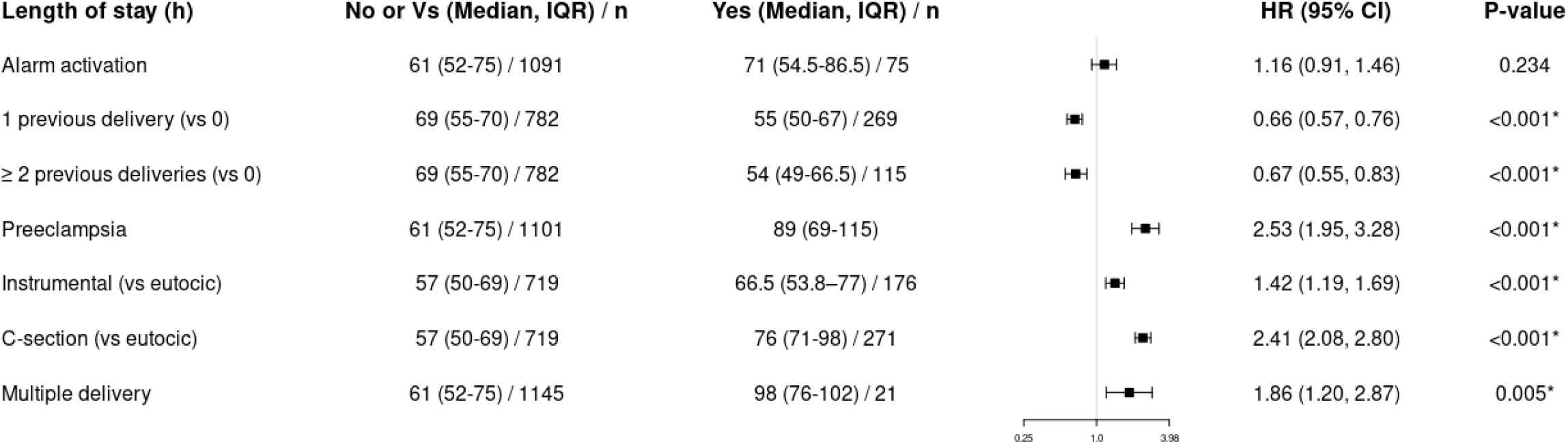
Forest plot of multivariate logistic analysis of the influence of MEWC activation, patient comorbidity and delivery on length of stay. We present the results as an odds ratio with a 95% confidence interval. Results less than 1, left of the y-axis, imply risk reduction. We accepted p < 0.05 as significant.

No significant p-value was rejected after calculating its q-value within the multiple comparability study (S4 File).

## Discussion

The first thing that draws attention to implementing the MEWC in our hospital is the data on sensitivity, specificity, positive, and negative predictive value of alarm activation. Our protocol’s sensitivity is very low (0.28), while specificity and negative predictive value are very high (0.94 and 0.99, respectively). For practical purposes, these results mean that we can be confident that patients without activation of the MEWC protocol alarm are doubtful to suffer a PFD. However, the cost was not to detect many patients who developed PFD (28 out of 39, 71.1% of total cases). Although we do not know the causes of such low sensitivity levels, we suspect that it is probably related to the patient’s monitoring period or the different parameters that form the protocol.

Our study monitored the patients for only 2 hours after delivery. However, other studies with better sensitivity results had a much more excellent follow-up. Singh et al. [13] published the first study to validate MEWC, carrying out a prospective observational study in 676 obstetric patients to detect morbidity between the 20th week of gestation and six months postpartum. 30% of the patients had an altered trigger. They defined a MEWC sensitivity of 0.98 and a specificity of 79%. The introduction of such an extended period of obstetric monitoring is probably impossible to implement in our center. Given our hospital’s structure, our patients go from the delivery room to conventional hospitalization rooms, where the monitoring is not comparable to that of a post-anesthesia recovery unit. Still, it makes us suspect that if we extend the monitoring period from admission to 24 hours after birth, we should reduce the number of false negatives.

On the other hand, the protocol modification by adding obstetric factors has also been able to modify the sensitivity and specificity of the protocol. Initially, we can think that the more complex a protocol is, the more difficult it is to generate false negatives, and the more secure its implementation is. However, this is not as true as it seems. Mhyre et al. [9] described that the simpler the warning criteria, the shorter the time for recognition, diagnosis, and treatment of women who are more at risk of developing obstetric complications. The more uncomplicated measures are more reliable, less vulnerable to human calculation errors, and have higher reproducibility. Mhyre et al. [9] insist on creating multidisciplinary teams that define their warning criteria for each center and review the evidence to date using these criteria9. In 2015 Edwards et al. [14] compared the predictive value of six different MEWS to identify severe sepsis in women with chorioamnionitis and informed of a sensitivity range of 40%-100% with a degree of 4%-97% specificity. The authors concluded that the MEWC tools with simpler designs tended to be more sensitive, while more complex ones tended to be more specific. By adding the obstetric items, we probably complicated the protocol and caused an increase in specificity and a decrease in sensitivity.

Likewise, a complex MEWC protocol that causes a high number of false positives is a severe limitation in its application since it can lead to professional fatigue due to the high number of false alarms15. Of course, no MEWC protocol is ideal, and we fully agree with Friedman et al. [15]. They argued that since each hospital has a unique environment, each hospital should develop its protocol and improve it, depending on its particular outcomes. We are currently in that phase, the improvement phase. Our study results encourage us to persevere in perfecting the protocol, either by increasing the time of monitoring the patient or by altering the variables to be monitored. However, further studies are necessary to validate and generalize MEWC principles.

## Conclusion

Our MEWC protocol presented low sensitivity and high specificity with a high negative predictive value, having many false-negative patients. Our study showed an association between alarm activation and PFD, reintervention rate, and critical care admission rate, with preeclampsia being the most related factor in all of them.

## Data Availability

All data has been self collected during fieldwork and is available for review if needed.

## Supporting information

**S1 File. Ethics committe approval document**. Original in Spanish. Date of Approval: 12th March 2018, Code: FJD-MEOWS-17-01.

**S2 File. Data collection form**

**S3 File. Original study databases**.

**S4 File. Statistical analysis**. Html format.

**S5 File. Statistical analysis raw R code**. R-Markdown format.

## Notes

### Competing Interest Statement

The authors have declared no competing interest.

### Funding Statement

no external funding was received.

### Author Declarations

Ethic Research Comittee at Hospital Fundacion Jimenez Diaz approved on the 24th October 2017

